# The Risk of Secondary Malignancies in Patients with Ovarian Cancer: A Systematic Review and Meta-Analysis

**DOI:** 10.1101/2020.04.07.20057190

**Authors:** Keyvan Heydari, Sahar Rismantab, Reza Alizadeh-Navaei, Amir Shamshirian, Nima Shadmehri, Parisa Lotfi, Alieh Zamani-Kiasari, Danial Shamshirian

## Abstract

This study was performed to systematically asses the risk of secondary malignancies in patients with ovarian cancer. We conducted a systematic search in PubMed, Web of Science, and Scopus databases up to September 2019 to find target studies. In this study, the overall SIR has been calculated with fixed/random-effects models. Sixteen cohort studies including 122715 ovarian cancer patients with 4458 secondary malignancies have been included in this meta-analysis. Combined SIRs showed an increased risk of secondary malignancies prevalence (SIR: 1.81, 95%CI 1.58-2.03). The most prevalence diagnosed malignancies were as follows: breast cancer (1.34, 95%CI 1.5-1.18), intestine (2.36, 95%CI 1.11-3.61), colorectal (1.73, 95%CI 1.44-2.02), pancreatic (1.42, 95%CI 1.13-1.71), cervical cancer (11.57, 95%CI 6.94-16.21), renal (1.43, 95%CI 1.11-1.74), bladder (2.13, 95%CI 1.77-2.50), leukemia (3.33, 95%CI 2.23-4.43), connective tissue (2.61, 95%CI 1.56-3.66), and thyroid (1.59, 95%CI 1.13-2.04). In regards to the results, various malignancies have a greater prevalence in patients with ovarian cancer in comparison to the general public. Corpus cancer, leukemia cancer, endometrium cancer, connective tissue malignancy, and bladder cancer are among the high risks in these patients and need to be considered for them. Hence, the survival rate of the patients can be increased through prevention and early diagnosis.

## Instruction

Ovarian cancer with 239000 diagnosed cases in 2012 was the 7th common cancer in females globally. This cancer has a 5 years survival rate in 30-50% of patients(Ferlay et al., 2015). In the United States, ovarian cancer is the second common and also the most deadly gynecologic cancer(Siegel et al., 2019). Advances in screening, surgery, and chemotherapy helped the improvement of survival rate in recent years. However, the risk of secondary malignancies, which endanger the lives of the patients, are causing concerns (Kim et al., 2012, Yoshino et al., 1985, Kim et al., 2013).

Women with ovarian cancer have a higher risk of secondary malignancies. Some of these malignancies are related to the side effects of chemotherapy/radiotherapy, like leukemia, bladder, and rectum. On the other hand, some of them like breast, colorectal, and stomach are probably related to genetic and environmental factors (Reimer et al., 1978, Storm and Ewertz, 1985, Kaldor et al., 1987, Levi et al., 1993, Travis et al., 1996a). In adults, malignant ovarian cancers mostly have epithelial cell origin and resulted in a lower survival rate (Baldwin et al., 2012). Furthermore, studies on malignant ovarian tumors in children and adults have shown that the major part of the ovarian tumors originated from germ cells (Brookfield et al., 2009). Tumors with germ cell origin are categorized as germinomas and nongerminomas. Moreover, the tumors with germ cell origin have been recognized as tumors with a very good prognosis. The 5 years and 10 years’ survival rate in them are 91.7 and 91.4 percent (Brookfield et al., 2009).

Our search hadn’t found any systematic review on risk assessment for secondary malignancies in female patients with ovarian cancers. Although, Zikry and his team had done a meta-analysis on the risk assessment of melanoma in patients with ovarian teratoma [13]. We conducted a systematic review and meta-analysis on risk assessment of occurring cancer in patients with ovarian cancer to find more comprehensive results.

## Material and Methods

### Information sources

This systematic review and meta-analysis had been conducted on cohort studies. We use PubMed, Scopus, and Web of Science databases with the publishing date of until September 2019.

### Search

The search in the PubMed, Scopus, and Web of Science databases had been done with the following keywords: ovarian neoplasm, incidence, second, secondary, after, and neoplasm in the title and abstract field.

In manual search part, after reference list added, the list of review references were selected.

### Eligibility Criteria

The criteria for selecting the papers were as the following:

- The study should have been cohorts and contain the patients with ovarian cancers.
- The study should declare the results in SIR or the SIR can be calculated with the presented data.
- The paper should have been in English or the English version was available and it should have been published before September 2019.
- There is no limit for the location of the study and all the appropriate studies have been considered without any regard to the location of the study.
- All the non-cohort studies including case series, case-control, and case reports have been omitted.
- All the animal and laboratorial studies have been omitted.

### Study selection

This stage of the study had been conducted in 3 parts. First, the duplicates were deleted by the Endnote software, then 2 researchers separately screened the remaining papers regarding the defined criteria. For those papers that have connections to our criteria but the title/abstract wasn’t enough the full texts were gathered and then the decision of the paper appropriation was made. If there were no agreement on an appropriation of a paper between the two primary researchers, the opinion of a third researcher was deciding. For this paper, we used the studies which have been done on risk assessments of second malignancies in ovarian cancer patients.

### Quality assessment

For quality assessment, the tool Newcastle-Ottawa Quality Assessment Form for Cohort Studies (NOS) has been used. The studies have been classified into 3 categories. Studies with a score of 1, 2, and 3 weak; studies with a score of 4, 5, 6 average; and the studies with a score of 7, 8, and 9 considered good.

### Data Extraction

Data extraction has been done by the checking full text of the papers. In each study, the information such as the author name, year of the publication, the average age of the patients, SIR of secondary malignancies, and the length of the follow up was extracted and stored.

### Statistical Analysis

The heterogeneity of the studies was screened by the I^2^ test and by the results of the test if the I^2^ were more than 50% the random/fixed effect model have been used to combine the results of the studies. The results were analyzed by the STATA v11 software and forest plot chart.

## Results

### Study selection process

With the conducted search 4288 papers were found and after omitting the duplicate papers they enter the screening stage, finally, after reviewing 39 full-text papers, 16 papers were used for the meta-analysis. The process of selecting data is available in the PRISMA Flowchart and chart 1 (Fig. 1).

**Figure 1.**
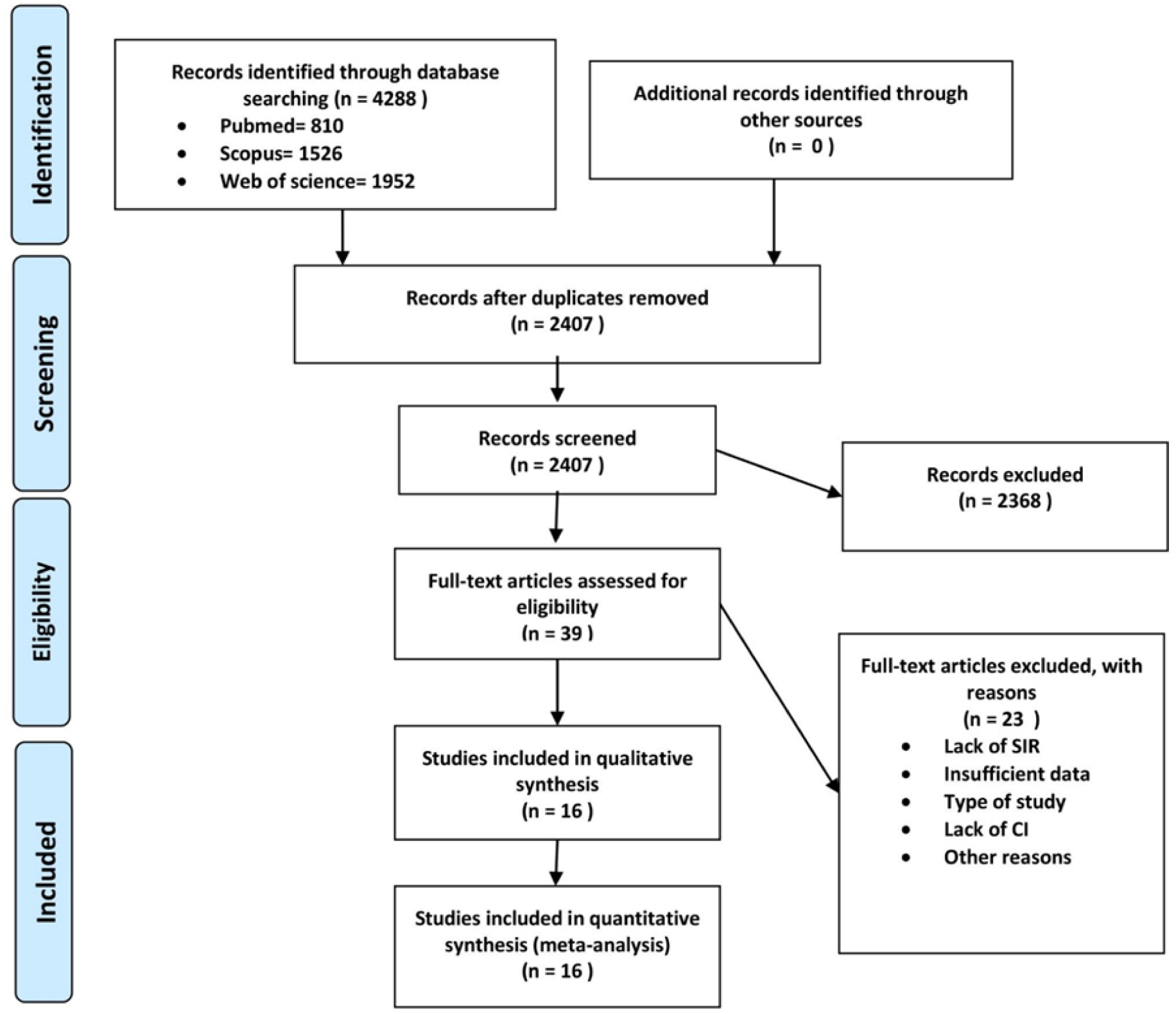
PRISMA flowchart for study selection process.

### Study characteristics

The variety of patients in the 16 cohort studies is from 130 to 32251 with the total number of patients add up to 122715 people. 4458 cases of secondary malignancies after the diagnosis of ovarian cancer had been reported. The location of the cohort studies is as following, 3 were from the US, 2 were from Sweden, 2 were from Switzerland, and 1 was from Germany, Australia, Israel, Netherland, Britain, turkey, Taiwan, and Finland. There is only one study that doesn’t clearly state the location of the study. The details of the cohort studies can be found in Table-1

### Quality assessment

In this research and by using the NOS checklist, we evaluated the methodological quality of the studies that we used in the meta-analysis. The result of the test is as following: 11 studies were good and 5 studies were average in quality.

### Risk assessment of secondary malignancies after ovarian cancer

Based on the cohort studies that have been used in the meta-analysis there are more than 40 types of secondary malignancies have been reported in patients with ovarian cancer.

Malignancies that been reported in only one study and were rare among ovarian cancer patients are as following: malignancies of the tongue, buccal, head and neck, gastrointestinal tract, larynx, uterus, ovary, vulva, eye, mesothelioma, bone, ALL, and CLL.

Cancers that been reported in more than one studies include: mouth/pharynx, esophagus, stomach, small intestine, colon, rectum, colorectal, liver, pancreas, digestive system, lung, respiratory system, breast, endometrium, cervix, corpus, genital, renal, bladder, melanoma, skin (excluding melanoma), urinary tract, brain, central nervous system, thyroid, endocrine glands, connective tissue, Hodgkin’s lymphoma, non-Hodgkin’s lymphoma, leukemia, and hematopoietic.

Only secondary malignancies that have been reported in more than one study entered the meta-analysis.

### All cancers

SIR of all cancers in ovarian cancer patients reported in 14 cohorts. Therefore, after meta-analysis on the reported SIRs, the total SIR was (1.81, 95%CI 1.58-2.03). Also, the severe statistical heterogeneity has been found in this cohorts (I^2^=94.1%). (Fig.2)

**Figure 2.**
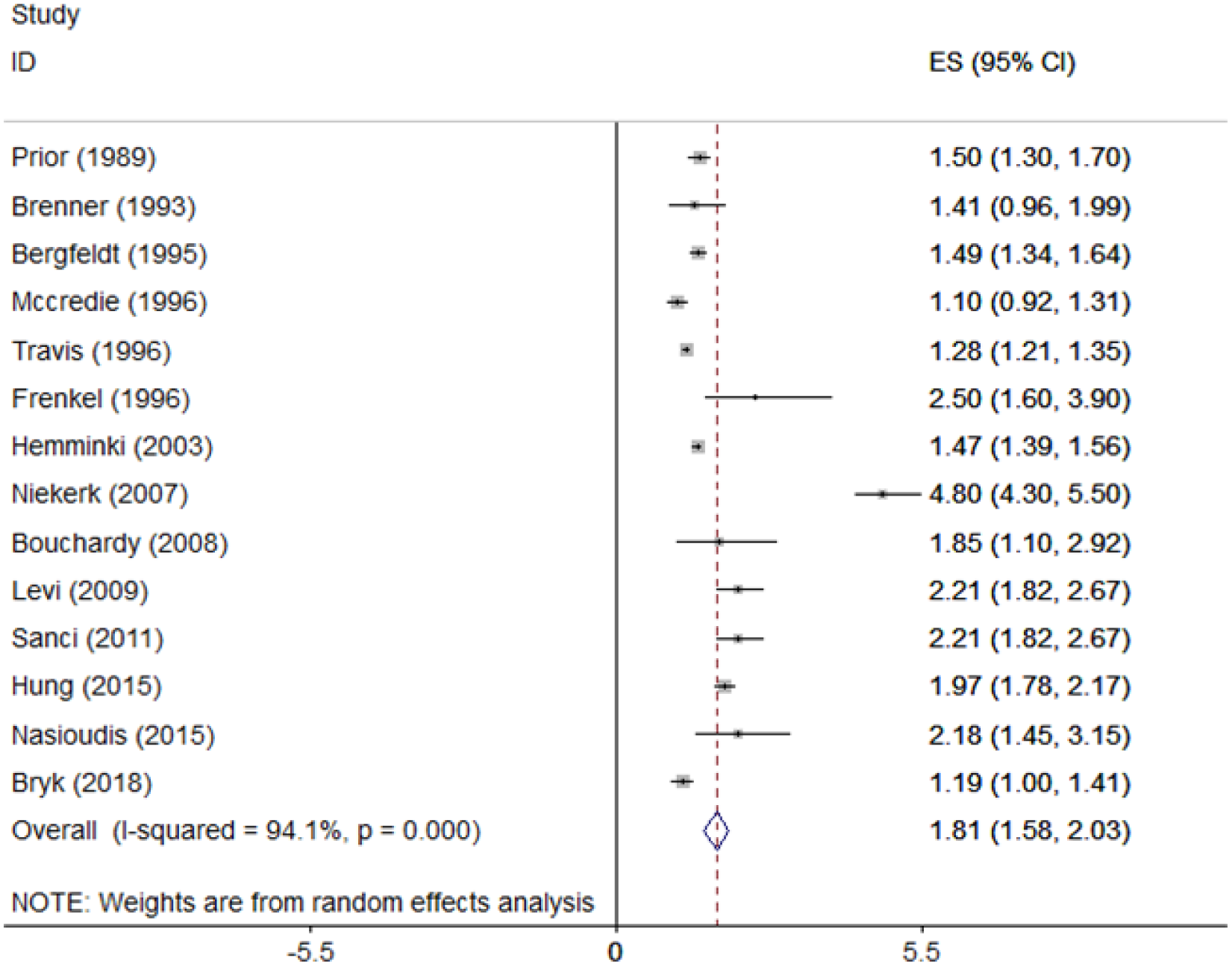
Forest plot for pooling SIR of All cancer.

### Breast cancer

There are 13 cohort studies with breast cancer report. The result of SIR meta-analysis on these data is (1.34, 95%C1 18-1.50) and the statistical heterogeneity is I^2^=58.2. (Fig.3)

**Figure 3.**
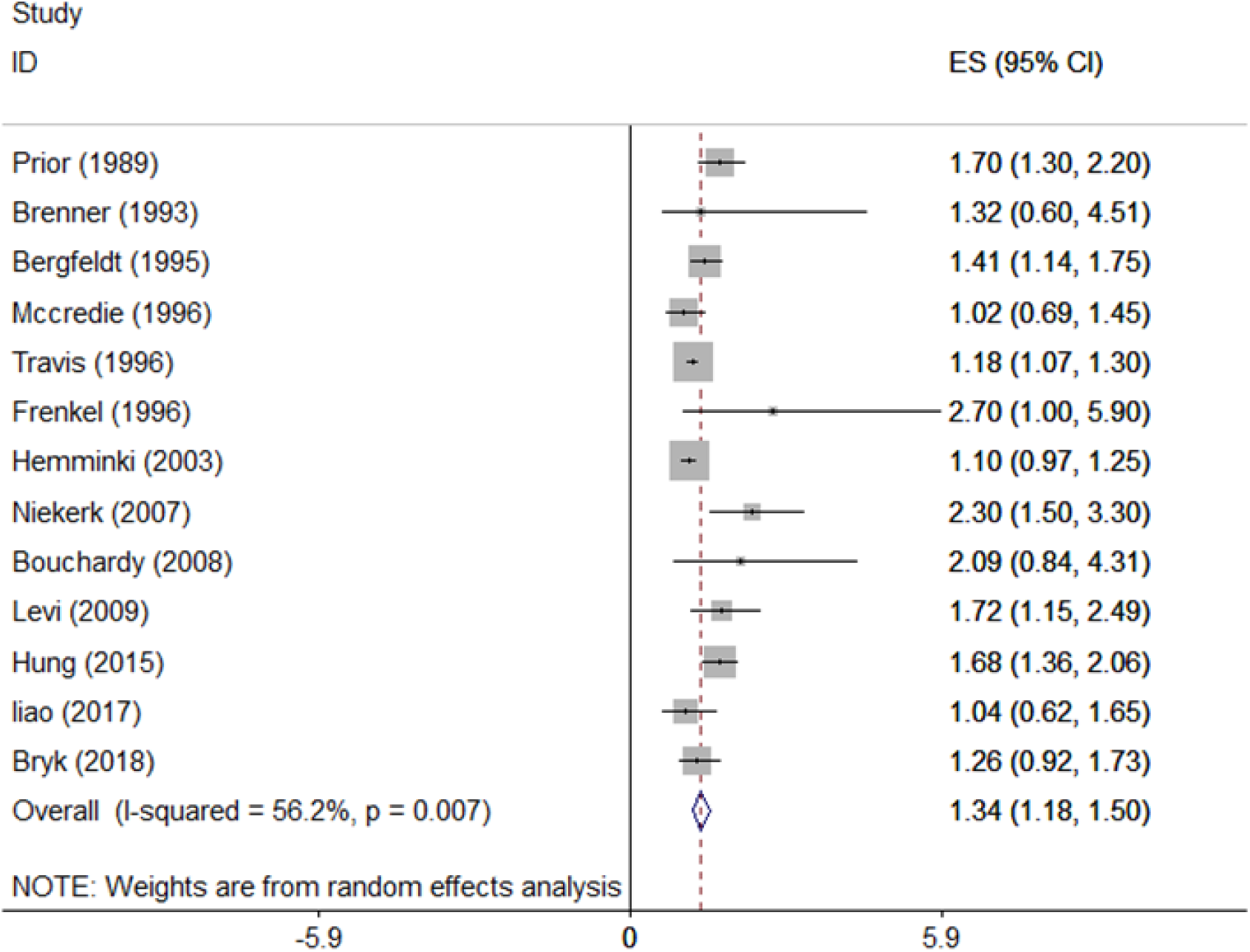
Forest plot for pooling SIR of Breast cancer.

### Digestive system

The mouth and pharynx malignancies have investigated in 4 studies. The total SIR in these studies was (1.7, 95%CI 0.31-3.09) and the heterogeneity with the I-squared index was very low. (Table 2)

**Table 1.** Characteristics of studies entered into the meta-analysis.

| Author (Year) | Region | Follow-up | person-years<br>Follow up | No. of<br>Participants | Total<br>Mean<br>age<br>(range) | No. of<br>Secondary<br>cancer | Quality<br>assessment<br>Score<br>(1to9) |
| --- | --- | --- | --- | --- | --- | --- | --- |
| Prior (1989)(Prior and Pope, 1989) | UK | 6.5 Y | 21446 | 7203 | - | 213 | 7 |
| Brenner (1993)(Brenner et al., 1993) | Germany | 3 Y | 19040 | 1745 | - | 32 | 7 |
| Bergfeldt (1995)(Bergfeldt et al., 1995) | Sweden | 6 Y | 27848 | 5060 | 59(1-80) | 346 | 7 |
| McCredie (1996)(McCredie et al., 1996) | Australia | 3.6 Y | 18570 | 5170 | 59.5 | 126 | 7 |
| Frenkel (1996)(Frenkel et al., 1996) | Israel | - | - | 364 | - | 21 | 7 |
| Travis (1996)(Travis et al., 1996b) | USA | 4.1 Y | 133098 | 32251 | 58.8 | 1296 | 8 |
| Weinberg (1999)(Weinberg et al., 1999) | USA | - | - | 28832 | - | 300 | 7 |
| Hemminki (2003)(Hemminki et al., 2003) | Sweden | - | 156877 | 19994 | 61* | 1104 | 8 |
| Niekerk (2007)(van Niekerk et al., 2007) | Netherland | - | - | 5366 | - | 244 | 5 |
| Bouchardy (2008)(Bouchardy et al., 2008) | Switzerland | 8.8 | - | 130 | 50.7(17-92) | 18 | 8 |
| Levi (2009)(Levi et al., 2009) | Switzerland | - | - | 304 | - | 31 | 5 |
| Sanci (2011)(Sanci et al., 2011) | Turkey | 96.5 M | - | 870 | 47(19-79) | 110 | 8 |
| Nasioudis (2015)(Nasioudis et al., 2015) | USA | - | - | 806 | 15.4 | 28 | 6 |
| Hung (2015)(Hung et al., 2015) | Taiwan | 3.48 Y | 56214 | 12127 | 50(42-61)* | 407 | 7 |
| Liao (2017)(Liao et al., 2017) | UC | 13.9 | - | 1507 | 22(0-86) | 45 | 6 |
| Bryk (2018)(Bryk et al., 2018) | Finland | - | - | 986 | - | 137 | 5 |

\*Median, Y= Year, M= Month, UC= Unclear

**Table 2.**
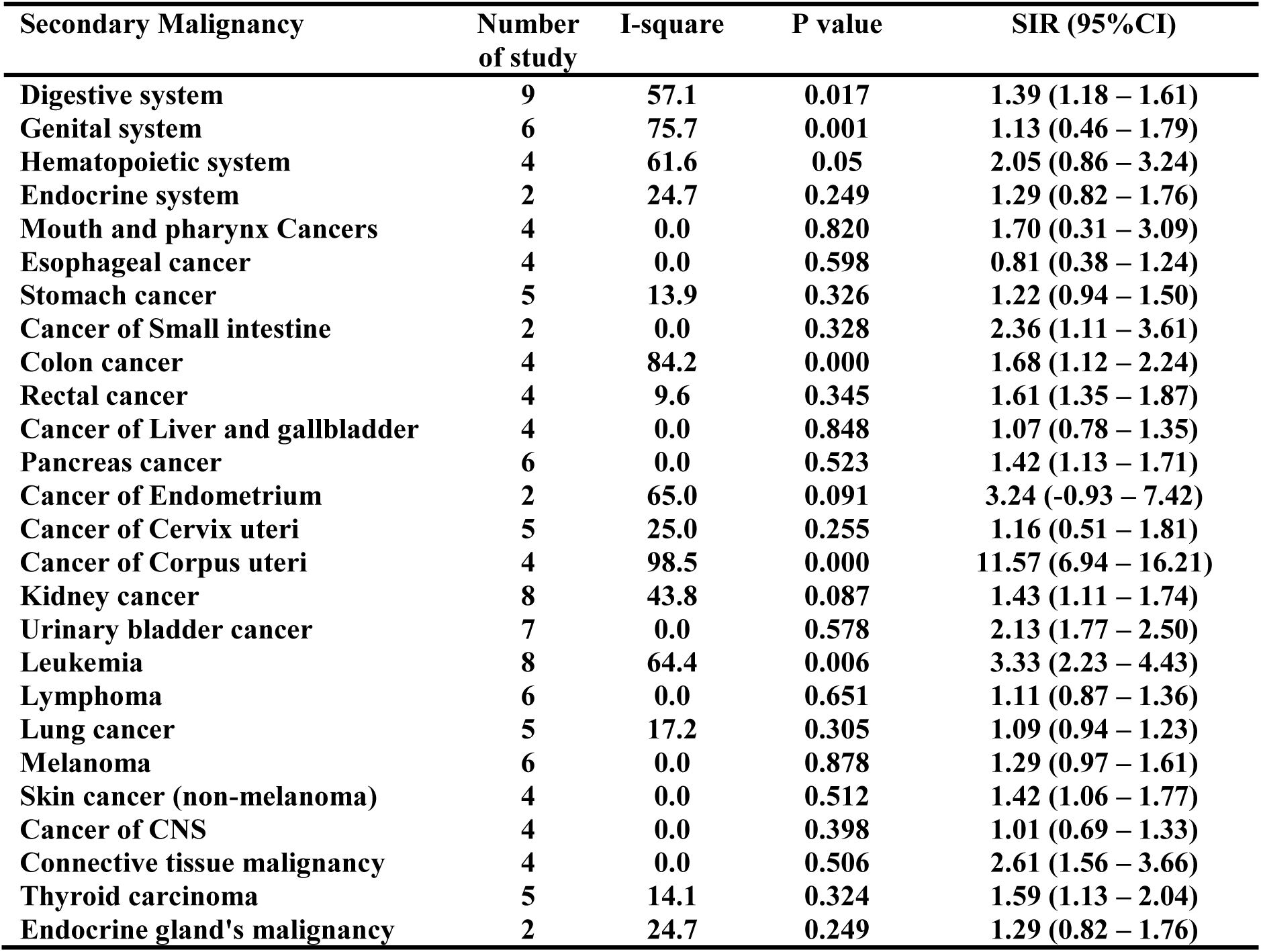
Summarized Pooled SIRs of Considered Secondary malignancies.

The esophagus and stomach secondary malignancies were investigated in 4 and 5 studies. The result of the meta-analysis for them is (0.81, 95%CI 0.38-1.24) and (1.22, 95%CI 0.94-1.50). The I^2^ index of these 2 cancers was 0% and 13.9%. (Table 2)

Small intestine cancer has been reported in 2 cohort studies. The meta-analysis of the data was (2.36, 95%CI 1.11-3.61) and the statistical heterogeneity was very low (I^2^=0%). (Table 2)

Cancers of the large intestine have been reported separately in different studies. SIR related to the colon, rectum, and colorectal were reported in 4, 4, and 11 studies; and the result for these cancers are respectively as following: colon (1.68, 95%CI 1.12-2.24) (I^2^=84.2%), rectum (1.61, 95%CI 1.35-1.87) (I2=9.6%), and colorectal (1.73, 95%CI 1.44-2.02) (I^2^=78.7). (Fig.4) (Table 2)

SIR of the liver and pancreas cancer in women with ovarian cancer have been reported respectively in 4 and 6 studies. After conducting the meta-analysis the overall SIR of the liver cancer was (1.07, 95%CI 0.78-1.35) and the meaningful heterogeneity wasn’t observed (I^2^=0%). The pooled SIR for pancreatic cancer is (1.42, 95%CI 1.13-1.71) and the heterogeneity is very low (I^2^=0%). (Table 2)

Digestive system cancers have been investigated in 9 studies with a total SIR of (1.39, 95%CI 1.18-1.61) and the heterogeneity of the studies which included in the meta-analysis is I^2^=57.1%. (Table 2)

**Figure 4.**
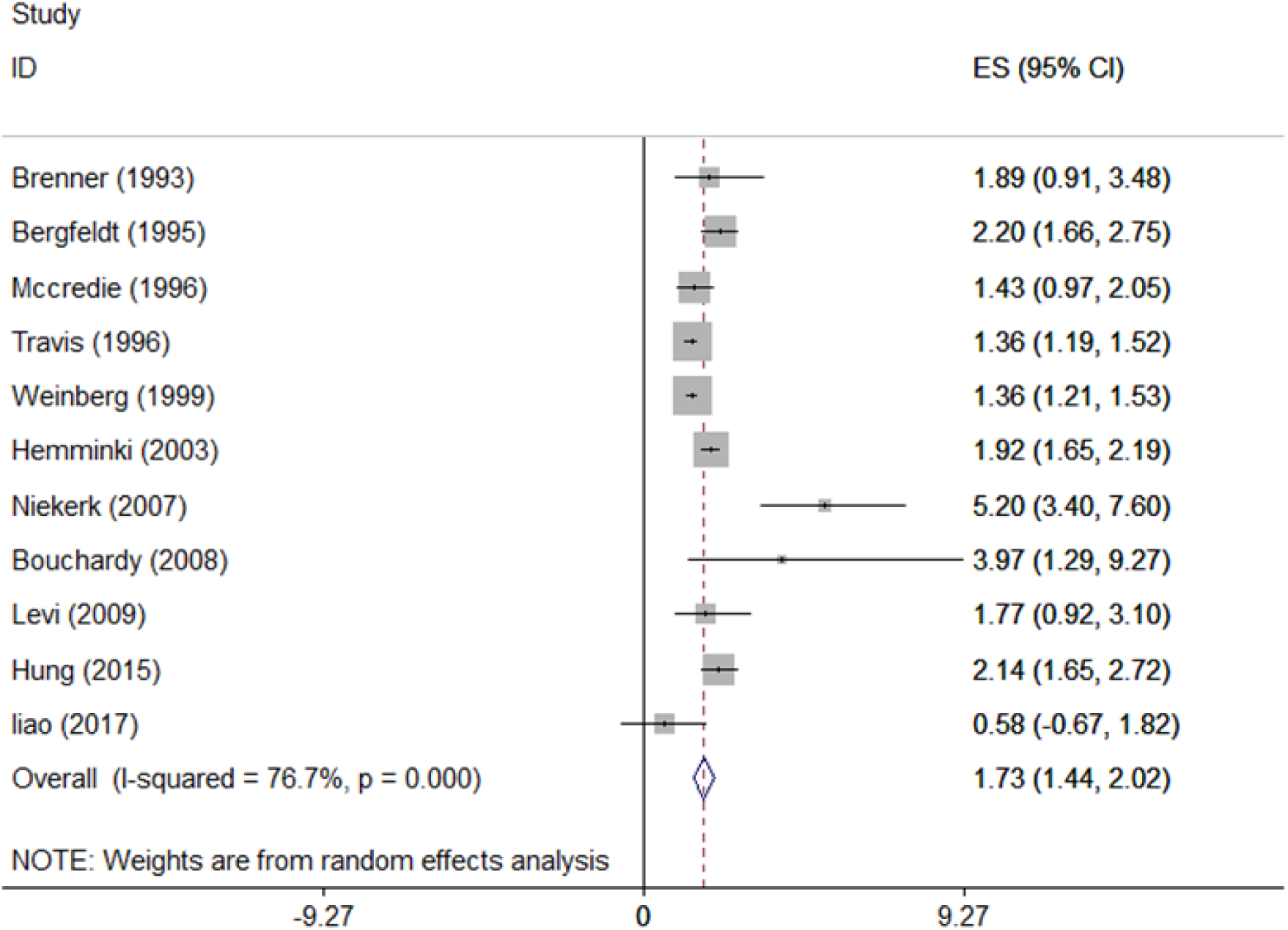
Forest plot for pooling SIR of Colorectal.

### Genital system

The endometrium, cervix, and corpus cancers are represented in 2, 5, and 4 studies. In this meta-analysis, the overall SIR of these cancers is (3.24, 95%CI −0.93-7.42), (1.16, 95%CI 0.51-1.81), (11.57, 95%CI 6.94-16.21). The respectively heterogeneity is (I^2^=65%), (I^2^=25%), and (I^2^=98.5%). (Table 2)

The SIR of women genital cancer in ovarian cancer patients has published in 6 studies and the meta-analysis of those data resulted in (1.13, 95%CI 0.46-1.79) with severe heterogeneity of (I^2^=75.7%). (Table 2)

### Urinary system

The SIR of renal and bladder cancer was reported in 8 and 7 cohort studies. The meta-analysis of the SIRs is as follows: (1.43, 95%CI 1.11-1.74) for renal and (2.13, 95%CI 1.77-2.50) for the bladder. The heterogeneity for the two are (I^2^=43.8%) and (I^2^=0.0%). (Table 2)

The data for urinary system malignancies were extracted from 10 studies. The result of meta-analysis for that is (1.84, 95%CI 1.55-2.13) with the heterogeneity of (I2=3.7%). (Table 2 and Fig. 5)

**Figure 5.**
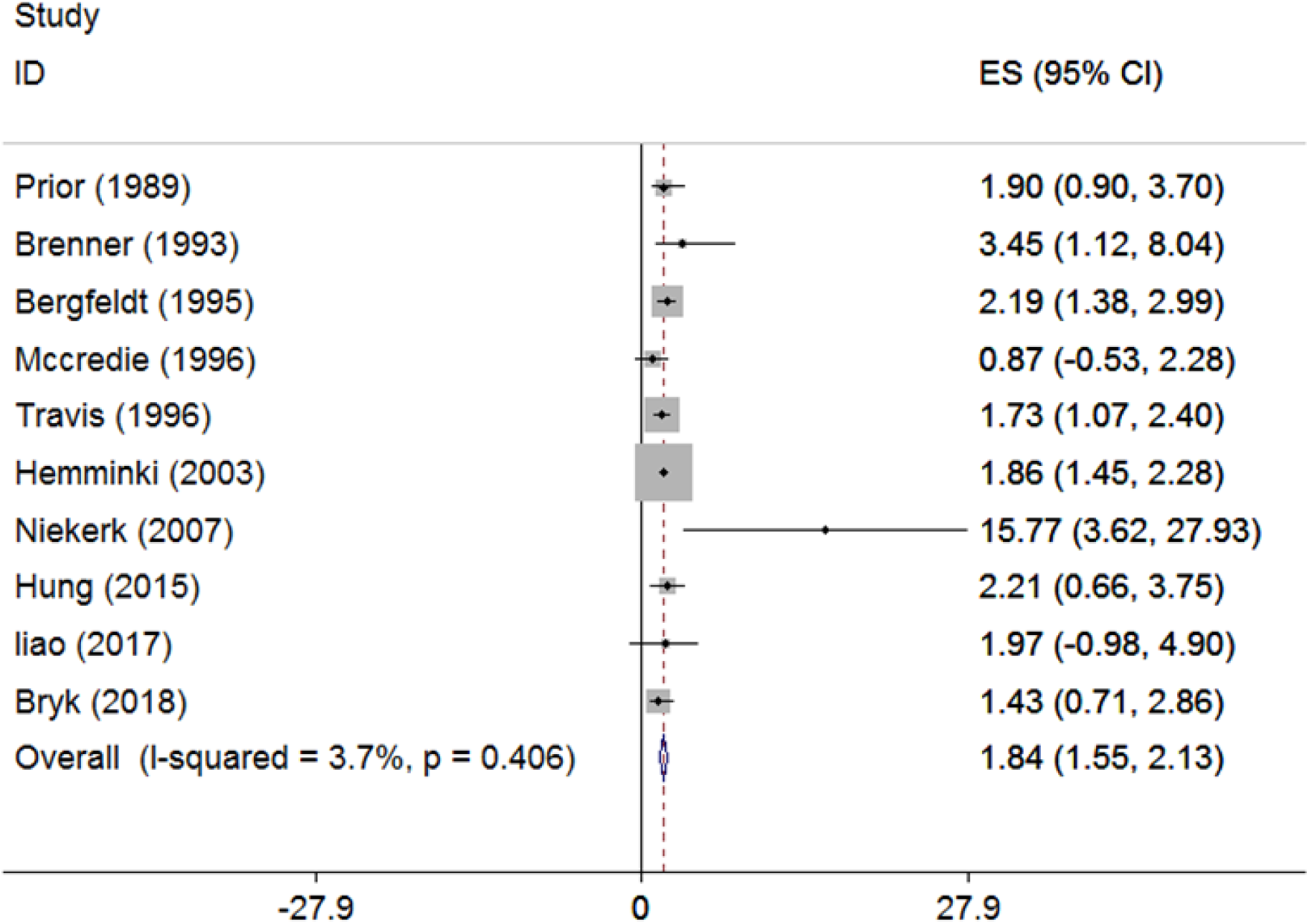
Forest plot for pooling SIR of Urinary system.

### Hematopoietic system

The occurrence of leukemia in ovarian cancer patients in contrast to the public was reported in 8 studies. After the meta-analysis on the reported data, the total SIR is (3.33, 95%CI 2.23-4.43) and the heterogeneity is (I^2^=64.4%). (Table 2)

The lymphoma in patients with ovarian cancer in contrast to the public was published in 6 cohorts. The sheer SIR is (1.11, 95%CI 0.87-1.36) and the heterogeneity is (I^2^=0.0%). (Table 2)

The data for hematopoietic system cancers were presented in 4 cohorts. With the meta-analysis done on the data the overall SIR is (2.05, 95%CI 0.86-3.24) and the heterogeneity is high (I^2^=61.6%). (Table 2)

### Respiratory system

The lung cancer incidents in ovarian cancer patients were investigated in 5 cohort studies. The total SIR is (1.09, 95%CI 0.94-1.23) with the heterogeneity of (I^2^=17.2%). (Table 2)

The existence of respiratory system cancers was reported in 3 cohort studies. After the meta-analysis on these data, the SIR is (1.23, 95%CI 0.43-2.02) and the statistical heterogeneity is (I^2^=50.3%). (Table 2)

### Skin cancers

SIR for melanoma and non-melanoma skin cancer were investigated in 6 and 4 studies. A meta-analysis on those data resulted in (1.29, 95%CI 0.97-1.61) and (1.42, 95%CI 1.06-1.77) and the heterogeneity was very low with (I^2^=0.0%) for both types of cancers. (Table 2)

### Central nervous system

There were 4 studies with the data regarding the brain and central nervous system. The overall SIR of those data resulted in (1.01, 95%CI 0.69-1.33) with statistical heterogeneity of (I^2^=0.0%). (Table 2)

### Endocrine system

SIR for the thyroid and malignancies of the endocrine system were reported in 5 and 2 cohort studies. The combined SIRs for them are (1.59, 95%CI 1.13-2.04) and (1.29, 95%CI 0.82-1.76). Additionally, the statistical heterogeneity of the data is (I^2^=14.1%) and (I^2^=24.7%). (Table 2)

## Discussion

In this systematic review and meta-analysis, we gather and extracted the data of secondary malignancies in patients with different ovarian tumors from 16 studies. Regarding the extracted data, it is understood that the risk of all secondary malignancies is high in these patients. The result of this research has shown that the incidents of the breast, small intestine, colorectal, pancreas, corpus, renal, bladder, leukemia, connective tissue, and thyroid cancer is higher in ovarian cancer patients in contrast to the public. Among these malignancies, the corpus cancer and leukemia, with SIR higher than 3, are more critical. Various factors can contribute to secondary malignancy incidents in people who are under treatment for ovarian cancer. One of the most important factors is genetic. Therefore, there are many investigations on the genetic roles in different cancers. One of the mutations which can cause ovarian cancer or increase the chance of the incident is breast cancer type 1 or 2 susceptibility genes (BRAC1, BRAC2). Hence, by different studies, it has been proven that these genes are involved in malignancies such as breast cancer (King et al., 2003, Gershoni-Baruch et al., 2000, Hodgson et al., 1999), endometrium cancer (Thompson and Easton, 2002, Moslehi et al., 2000, Segev et al., 2013), pancreatic (Thompson and Easton, 2002, Consortium, 1999, Iqbal et al., 2012), stomach, gallbladder (Consortium, 1999, Moran et al., 2012), and colorectal cancer. Besides, in a meta-analysis done by Oh and colleges which studied 14 case-control studies, it has been reported that pathogenic variants of the BRCA gene are connected to the risk of colorectal cancer (Oh et al., 2018). Also, the mutation of BRCA gene is associated with ovarian cancer (King et al., 2003, Gershoni-Baruch et al., 2000, Hodgson et al., 1999) and because after the treatment of patients with ovarian cancer this mutation is still present in their genome it can cause other types of cancers; so, there is an increasing chance for the occurrence of cancers in which this gene has an etiologic role. In our study, the higher the risk of incidents of breast, endometrium, pancreas, stomach, gallbladder, and colorectal have been observed. In a review paper by Denlinger et al. (Denlinger and Weinberg, 2009) in which he looks at ovarian cancer as a risk factor for colorectal cancer, he concluded that the occurrence of ovarian cancer will indeed higher the risk of colorectal cancer and that finding is in agreement on what we have found in this study.

## Conclusion

Regarding the results, in patients with ovarian cancer the wide variety of malignancies happen more than the normal public. Corpus, leukemia, endometrium, connective tissue and bladder cancers having a higher risk of occurrence in patients with ovarian cancer. Therefore, it is better to put these malignancies in the screening test for ovarian cancer patients so with better prevention and diagnose, the survival rate of the patients improves.

## Data Availability

The data that support the findings of this study are openly available in data bases mentioned in the search strategy.

## Acknowledgment

The authors would like to thank the Student Research Committee of Mazandaran University of Medical Sciences for supporting this project (Project No: 6594).

## Conflict of interest

The authors have no conflicts of interest to report.

## Notes

### Competing Interest Statement

The authors have declared no competing interest.

### Funding Statement

None

